# Wastewater-based surveillance is an efficient monitoring tool for tracking influenza A virus in the community

**DOI:** 10.1101/2023.08.28.23294723

**Authors:** Lehto Kirsi-Maarit, Hyder Rafiqul, Länsivaara Annika, Luomala Oskari, Lipponen Anssi, Anna-Maria Hokajärvi, Heikinheimo Annamari, Pitkänen Tarja, Oikarinen Sami, WastPan Study Group

**Affiliations:** Tampere University, Faculty of Medicine and Health Technology, Arvo Ylpön katu 34, 33520 Tampere, Finland; Finnish Institute for Health and Welfare, THL, Department of Health Security, Neulaniementie 4, 70210 Kuopio, Finland; Department of Food Hygiene and Environmental Health, Faculty of Veterinary Medicine, Agnes Sjöbergin katu 2, FI00014 University of Helsinki, Finland; Finnish Food Authority, Ruokavirasto, Alvar Aallon katu 5, 60100 Seinäjoki, Finland

**Author notes:** Correspondence: Sami Oikarinen.

## Abstract

Around the world, influenza A virus has caused severe pandemics, and the risk of future pandemics remains high. Currently, influenza A virus surveillance is based on the clinical diagnosis and reporting of disease cases. In this study, we apply wastewater-based surveillance to monitor the incidence of the influenza A virus at the population level. We report incidence data of the influenza A virus in 10 wastewater treatment plant catchment areas covering 40% of the Finnish population. Altogether, 141 monthly composite influent wastewater samples (collected between April 2020 and May 2021) were analysed from supernatant fraction using influenza A virus specific RT-qPCR method. During the study period, an influenza A virus epidemic occurred in two waves in Finland. This study shows that the influenza A virus can be detected from the supernatant fraction of 24 h composite influent wastewater samples. The influenza A virus gene copy number in wastewater correlated with the number of confirmed disease cases in the Finnish National Infectious Diseases Register. The Kendall’s τ correlation strength was over 0.600 in all wastewater treatment plants, save for one, which was 0.459. The strongest correlations were 0.761 and 0.833. Wastewater-based surveillance of the influenza A virus is an unbiased method and cost-efficiently reflects the circulation of the virus in the entire population. Thus, wastewater monitoring complements the available, but often too sparse, information from individual testing and improves health care and public health preparedness for influenza A virus pandemics.

## Introduction

The emergence of viral outbreaks, local epidemics and further global pandemics are difficult to predict. Pandemics because of viruses cause uncontrollable effects on society worldwide, as has been seen with SARS-CoV-2. Classical epidemiology and management systems are based on the diagnostics of samples obtained from patients with clinical symptoms (Dalmau Llorca et al., 2022). This approach fails to detect epidemics threatening public health at an early stage because all infected persons are not diagnosed, and because of the time lag between the infection, clinical symptoms, and the diagnosis. These infected but undiagnosed cases spread the virus, and epidemics may be established prior to the recognition by health care professionals. Therefore, clinical testing has failed to notice the epidemic in the initiation phase (Robins et al., 2022).

Municipal wastewater reflects comprehensive unbiased health information of the entire community; it represents an up-to-date snapshot of the status of the public health in the wastewater treatment plant (WWTP) area. Untreated influent wastewater is a versatile sample matrix from the population because it consists of various human secretions, such as faeces, urine, vomit and water used for showering and brushing teeth. Wastewater is, therefore, a rich source of various biomarkers for monitoring public health. Wastewater-based surveillance (WBS) analyses wastewater to determine the exposure to pathogens at the population level (Safford et al., 2022); it has been utilised for the monitoring of illegal drugs, poliovirus and, lately, for monitoring COVID-19 pandemic (Duintjer Tebbens et al., 2017; Hovi et al., 2012; Sulej-Suchomska et al., 2020; Medema et al., 2020). SARS-CoV-2 has been detected in wastewater prior to an outbreak in a certain area, showing that the WBS can be used as an early warning system for viral epidemics. Similarly, WBS can be used for monitoring SARS-CoV-2 pandemic outbreaks because the SARS-CoV-2 gene copy (GC) number in wastewater correlates with and predicts changes in the prevalence of clinical cases (Medema et al., 2020).

Influenza A virus (IAV) has caused five severe pandemics in the world over the past 100 years. The number of deaths has been estimated to be at least 53 million worldwide due to pandemics caused by IAV variants H1N1, H2N2 and H3N2. IAV is one of the most probable viruses to cause severe worldwide pandemics also in the future (Khanna et al., 2012; Palese, 2004), and here, wastewater is a potentially viable matrix for population-level surveillance of IAV epidemics. First, IAV RNA was detected in 47% of stool samples from hospitalised adults with IAV infection (Chan et al., 2011). In addition, the association of IAV in wastewater and disease cases has been studied in three epidemics: one city and two university campus areas (Mercier et al., 2022; Wolfe et al., 2022).

In the present study, we report the monitoring results of IAV in the wastewater of 10 WWTPs in Finland between April 2021 and May 2022 in conjunction with the laboratory-confirmed IAV cases in the sewershed catchment area. We correlate the IAV GC number in wastewater with the number of reported influenza A cases in the Finnish National Infectious Diseases Register (NIDR).

## Materials and methods

### Sample material

#### 2.1. WastPan study

A WastPan project developed wastewater-based preparedness tools for environmental surveillance of infectious agents and antimicrobial resistance genes circulating in communities. Furthermore, it developed a platform used to convey the observed alarming microbial trends to policymakers and citizens. The WastPan project was implemented by a consortium that consists of the Finnish Institute for Health and Welfare (THL), Tampere University (TAU) and the University of Helsinki (UH) in Finland between the years 2020 and 2023.

The wastewater samples in the WastPan project were collected from 10 WWTP in Finland. The main selection criterion of the WWTPs was geographical coverage, and both larger and smaller cities were selected (between 30,500 and 860,000 inhabitants). The selected wastewater treatment plants were a representative fraction of a total of 28 WWTPs that have been involved in the national wastewater studies in Finland for illicit drug monitoring (Kankaanpää et al., 2016) and COVID-19 surveillance (Tiwari et al., 2022). Further, part of the WWTPs were also included in national poliovirus surveillance (Pöyry et al., 1988). The WWTPs are located in Helsinki (Viikinmäki), Espoo (Suomenoja), Turku (Kakolanmäki), Tampere (Viinikanlahti), Seinäjoki (Seinäjoen keskuspuhdistamo), Pietarsaari (Alheda), Lappeenranta (Toikansuo), Kuopio (Lehtoniemi), Oulu (Taskila) and Rovaniemi (Alakorkalo). These WWTPs represent about 2.2 million inhabitants and 40% of the Finnish population (Fig 1). The 24 h composite samples from untreated influent wastewater were collected monthly between February 2021 and February 2023. A fraction (∼ 1 litre) of the samples were shipped in cool boxes to three participating laboratories in THL, TAU and UH. The inside temperature of the cool boxes was monitored during the shipment using temperature data loggers (Winlog Basic, Ebro). Upon arrival to the laboratory, the temperature records from the data loggers were downloaded, the arrival time was recorded, sample temperatures were measured, and the samples were aliquoted. Prior to analysis, the samples were stored at 5 °C, and the excess sample fractions were stored in the sample banks at -80 °C freezers for later use in the analysis of monitoring targets with public health relevance.

**Figure 1.**
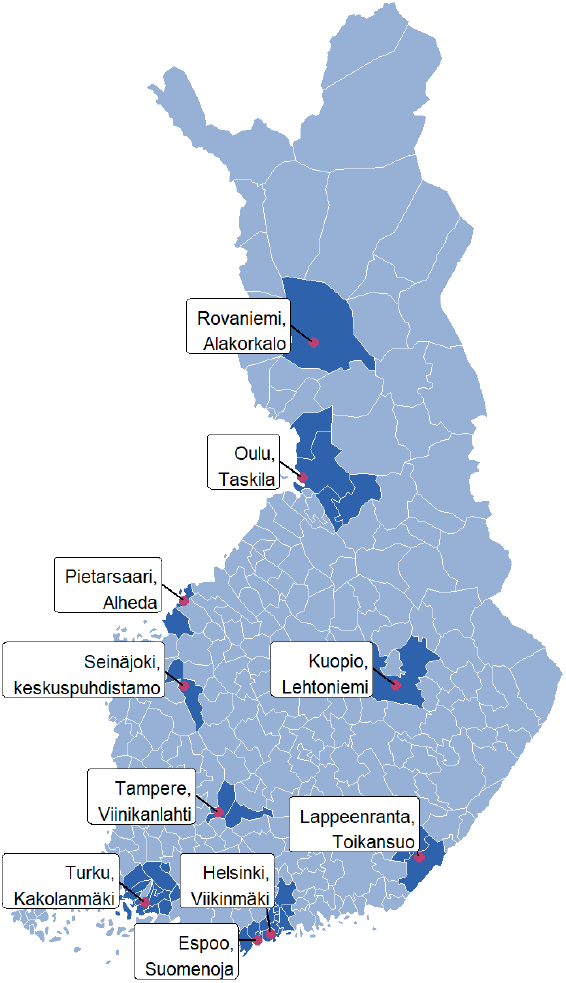
The WastPan study population consists of 10 WWTPs, which represent about 2.2 million inhabitants and 40% of the Finnish population (Municipal boundaries: Statistics Finland 2023).

### IAV quantification method

For virus analysis, an aliquot of 100 ml was processed without freezing within 24–48 h of sample collection (Hokajärvi et al., 2021). The samples were centrifuged at 4654 g for 30 min without a break to remove interfering particles. After the initial centrifugation step, the supernatant was concentrated with centrifugation at 3500 g for 15 min using a Centricon® Plus-70 centrifugal ultrafilter with a cut-off of 100 kDa (Millipore, Cork, Ireland). Then, the concentrated sample was collected from the filter by centrifugation at 1000 g for 2 min. Nucleic acids were extracted from the samples immediately after concentration using the PerkinElmer Chemagic Viral DNA/RNA 300 (Perkin Elmer, USA) extraction kit. The extraction was done according to the manufacturers’ protocol using an elution volume of 100 µl. The extracted nucleic acids were stored in a -80 °C freezer.

Reverse transcription-quantitative polymerase chain reaction (RT-qPCR) assay was used for quantifying IAV. The primers and probes recommended by the WHO (WHO, 2021) were procured by Merck (Finland). RT-PCR reaction was performed with TaqMan™ Fast Virus 1-Step Master Mix (Thermo fisher/Applied Biosystems™) in 25 µl volume, including 5 µl of template, 900 mM assay specific forward (InfA Forward 3’-GACCRATCCTGTCACCTCTGAC-5’) and reverse primers (InfA Reverse 3’-AGGGCATTYTGGACAAAKCGTCTA-5’), 100 nM probe (InfA Probe 3’-6-FAM-TGCAGTCCTCGCTCACTGGGCACG-TAMRA-5’) and 6.25 µl TaqMan Fast Virus 1-step MasterMix. To quantify the IAV in wastewater, a standard curve of the dilution series of control RNA with a known quantity (in-house pET3a-1a, 2.83* 10^1^–10^5^ GC per µl) was generated for all runs (Supplement material 1). PCR negative (RNase-free water) and nucleic acid extraction controls were included in every test run. The qPCR cycling conditions for the assay were used as reverse transcription at 50 °C for 5 mins, enzyme activation at 95 °C for 20 secs, 50 cycles of denaturation at 95 °C for 15 secs, annealing and extension at 58 °C for 1 min and then an infinite hold at 15 °C. Samples were run as duplicate in two runs using the QuantStudio 5 System (QuantStudio™ Design & Analysis Software v1.4.3), and the average of the gene copy (GC) numbers was calculated. A Ct value of <40 was used as a detection limit for positive test results. The limit of detection (LOD) of the IAV RT-qPCR method was evaluated with a dilution series of control RNA. The LOD for the IAV RT-qPCR was 4.8 copies per reaction. The performance of the RT-qPCR methods is shown in Supplementary Material Figure S1. The performance of the IAV RT-qPCR method was evaluated using an international quality control panel (QCMD 2019 influenza virus A and B RNA EQA Programme–influenza virus A); the method detected all samples correctly and did not cross-react with influenza B virus.

The IAV GC number in the wastewater was normalised to the daily WWTP flow by multiplying gene copies per m^3^ with the 24 h inflow of a WWTP (m^3^) and then dividing the results by the estimated population served at the WWTP, as previously described (Tiwari et al., 2022).

### Clinical data

The Finnish Institute for Health and Welfare maintains the NIDR, in which clinical laboratories report laboratory-confirmed IAV cases in a timely manner. The sum of 7 d IAV cases from NIDR was calculated for each WWTP catchment area. The NIDR data were adjusted to the WWTP catchment area as previously described (Tiwari et al. 2022). The sum of IAV cases from all cities in the WWTP catchment was calculated and then divided by the proportion of the estimated number of inhabitants in the WWTP catchment area. The number of IAV cases in the NIDR was adjusted according to the estimated number of inhabitants served by the WWTP divided by the total number of inhabitants in the municipalities served by each WWTP. The IAV GC in wastewater was correlated to the 7 d sum of IAV incidence per 100,000 persons in the NIDR.

### Statistical analysis

The null hypothesis was that IAV GC numbers in wastewater were not associated with the 7 d incidence rates of IAV in NIDR. None of the WWTP catchments data sets were normally distributed (Shapiro–Wilk normality test, *p* < 0.001). Therefore, the correlations between the IAV GC number in wastewater and clinical cases of IAV were evaluated using the Kendall rank correlation coefficient. Correlation coefficient 0 indicates no correlation, -1 indicates perfect negative, and +1 indicates perfect positive correlation. Virus gene copy numbers lower than the LOD were coded as 0.001, which was lower than any of the quantitated virus GC numbers in the analysed samples. All statistical analyses were done using IBM® SPSS® software, version 28.0.1.0 (142).

## Results

During the study period from April 2021 to May 2022, at least a few IAV cases were reported every month, except in August and September 2021 in Finland. IAV cases increased in 10 WWTPs in November 2021, with a small peak in December 2021. Few IAV cases were reported in the winter months of 2021 until a second wave of IAV cases began in April 2022, peaking in May 2022. The wave ended in July 2022 (Fig. 2).

**Figure 2.**
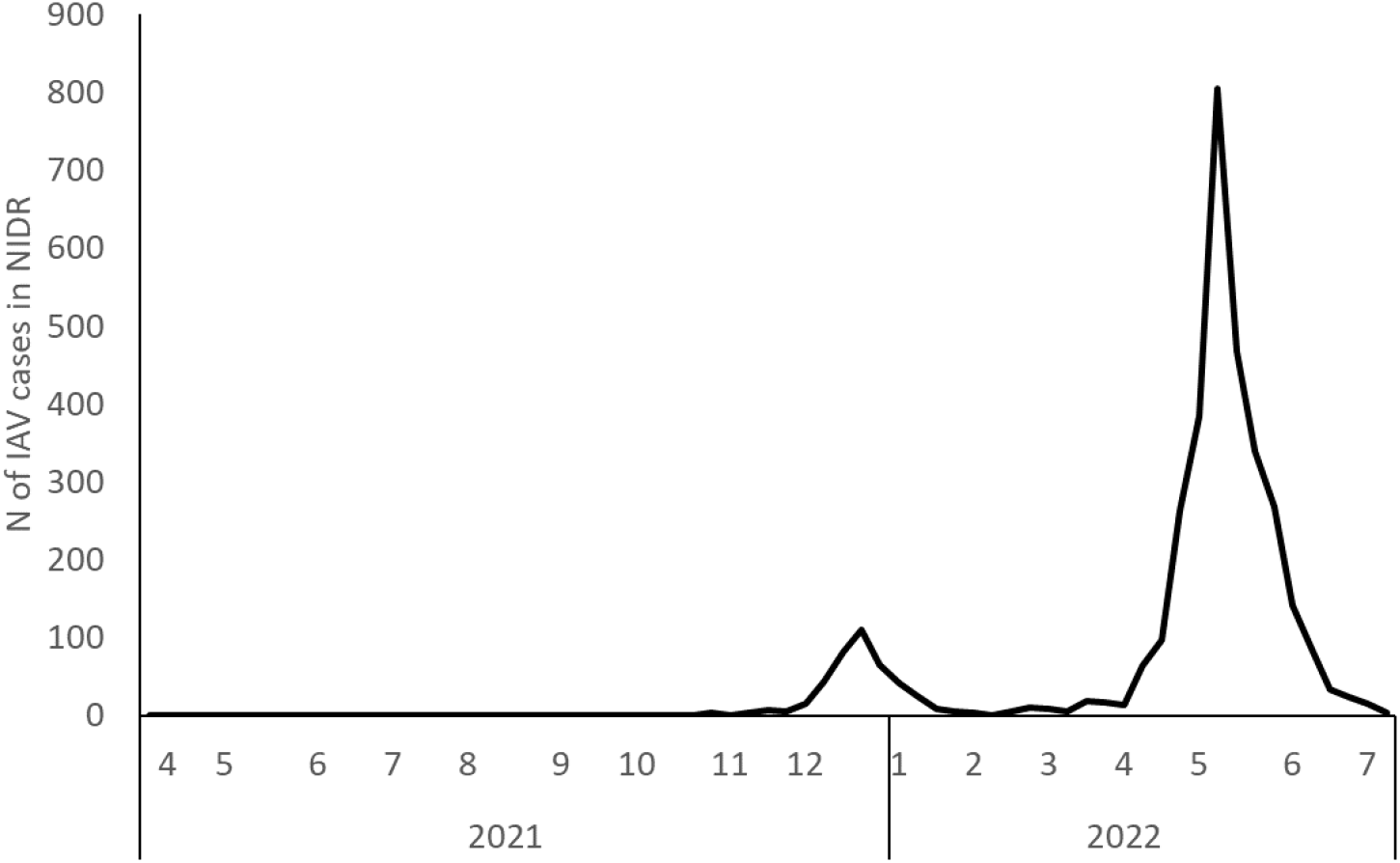
The total number of IAV cases in the NIDR in the 10 WWTP catchment areas in Finland during the study period.

Altogether, 141 monthly influent wastewater samples collected from 10 WWTPs were analysed. Most of the samples (98 samples) were collected when no IAV cases were reported in the corresponding catchment in the same week, and 43 samples were collected when at least one IAV case was diagnosed. During the weeks, when IAV cases were detected in the sewershed areas, the mean number of IAV cases was 17, median 6 cases and maximum 97 cases.

### IAV in wastewater qPCR

The IAV RNA was detected and quantified in 94.7% of the samples when the incidence of IAV was over 5 cases/100,000 persons/week. Furthermore, in the incidence group of one to five cases, 70.0% of the wastewater samples were positive for IAV. In the incidence group with less than one case, the IAV positivity of the wastewater samples decreased to 42.9%. The virus was detected in 11.2% of the samples when no cases of IAV were reported in the NIDR in the respective areas (Table 1). However, seven of these IAV-positive wastewater samples were collected one to two weeks apart from the IAV cases reported in the WWTP area, and another four samples were over 1 month apart from IAV cases in NIDR.

**Table 1.** The comparison between the reported IAV cases in NIDR and the number of wastewater samples above the LOD. Wastewater samples were grouped according to the incidence of IAV in the WWTP catchment area during the sampling week.

| Incidence<br>(IAV cases/100,000/week) | N of analysed<br>samples | N of IAV pos.<br>samples | Pos. samples<br>(%) |
| --- | --- | --- | --- |
| >10 | 10 | 9 | 90.0 |
| 5–10 | 9 | 9 | 100.0 |
| 1–5 | 10 | 7 | 70.0 |
| >0–1 | 14 | 6 | 42.9 |
| 0 | 98 | 11 | 11.2 |

The IAV GC number in wastewater was positively and significantly correlated to the reported clinical cases over the study period. However, the proportion between IAV GC number in wastewater and incidence in NIDR data showed a variation between the WWTPs (Fig. 3). The difference between WWTPs remained after the IAV gene copies in wastewater were normalised for population size and flow. Therefore, Kendall’s tau was calculated for individual WWTPs. The strongest correlation was observed with the sum of IAV incidence calculated 0–7 d after wastewater sample collection. The Kendall’s τ correlation strength was over 0.600 in all WWTPs, save for one (0.459). The strongest correlations were 0.761 and 0.833. In eight WWTPs, the results were statistically significant (Table 2).

**Figure 3.**
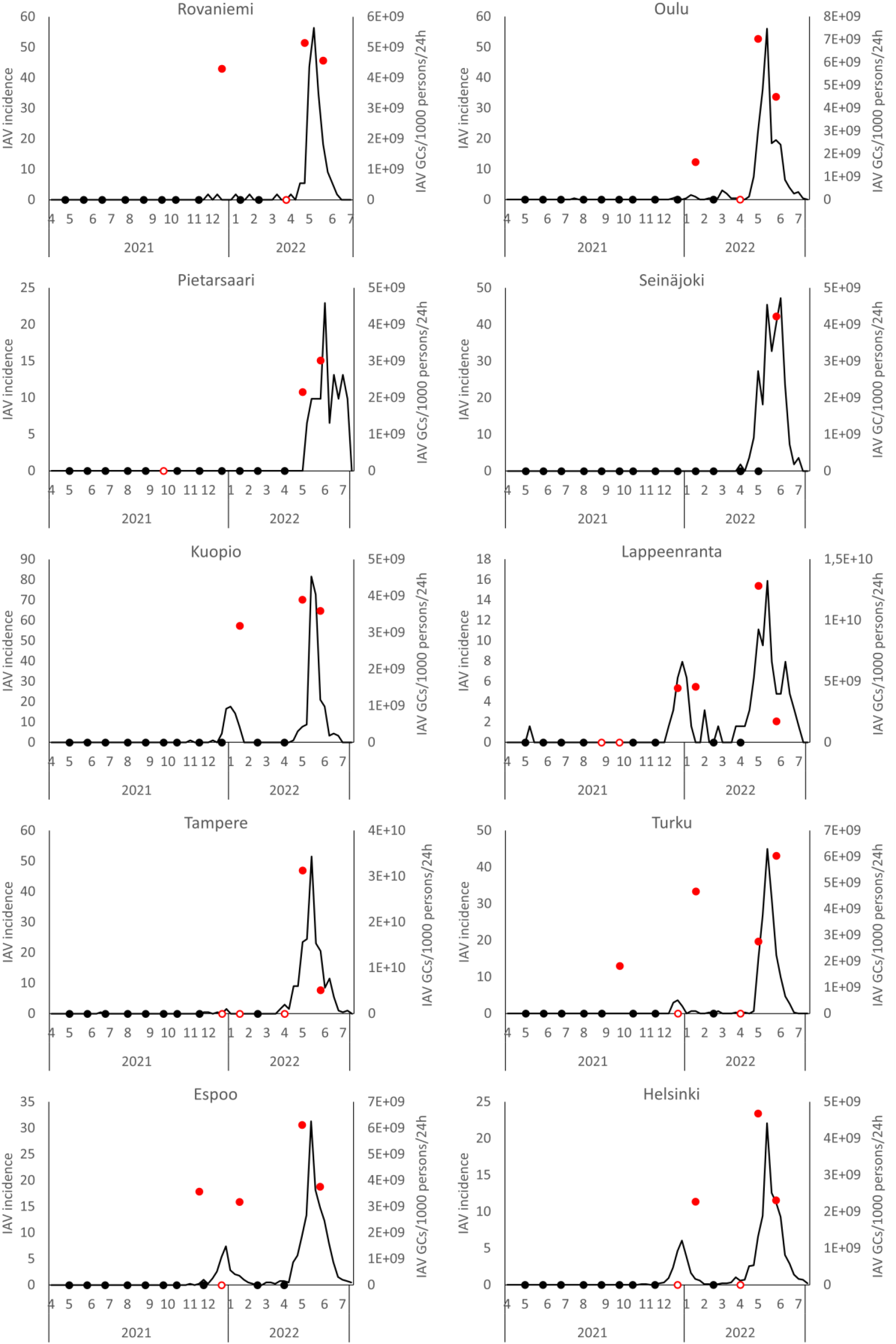
Influenza A GC number in wastewater correlates with the number of IAV cases reported in the NIDR across the time series. The results are shown for each WWTP. The black line shows the weekly incidence of influenza per 100,000 people in share of the population served by the wastewater network, and the red circles indicate the amount of the IAV GC per 1000 persons per 24 h in wastewater. The red rings indicate that influenza was detected in wastewater, but the concentration was below the limit of quantification, and the black circles indicate that IAV was not detected.

**Table 2.** The strength of the association between the IAV GC number in wastewater and clinical IAV NIDR data in each WWTP catchment. The table shows the number of analysed samples, Kendall’s τ and statistical significance for each WWTP. The wastewater data are correlated to NIDR data of the same collection week.

| WWTP | N of analysed samples | Kendall's $\tau$ | Significance |
| --- | --- | --- | --- |
| Rovaniemi | 14 | 0.747 | 0.027 |
| Pietarsaari | 14 | 0.601 | 0.261 |
| Turku | 14 | 0.699 | <0.001 |
| Kuopio | 14 | 0.833 | 0.001 |
| Seinäjoki | 14 | 0.610 | 0.270 |
| Espoo | 14 | 0.459 | 0.001 |
| Oulu | 14 | 0.761 | <0.001 |
| Lappeenranta | 14 | 0.605 | <0.001 |
| Helsinki | 15 | 0.730 | <0.001 |
| Tampere | 14 | 0.736 | 0.002 |

The sensitivity of RT-qPCR to detect the initiation of the IAV waves was analysed from 15 occasions, in which the incidence peak was at least five. Five occasions from the first wave were left out from the analysis because only sporadic IAV cases were reported in those WWTPs and evident continuous disease waves did not occur at these sites. The mean number of cases was 2, and the average was 2.2 at the initiation timepoints. The IAV was detected in wastewater a week before the reported IAV cases in two out of three samples; at the same week when first IAV cases were reported, one out of three samples were positive for IAV in wastewater. Further, a week after the initiation of IAV waves, three out of four samples, and two weeks after initiation, three out of four samples were IAV positive in wastewater (Table 3).

**Table 3.** The temporal correlation of the IAV in wastewater and initiation of the IAV waves in 10 WWTPs. The samples were clustered weekly according to the time from the first reported IAV case in time series in WWTPs (0 point). Twenty-four-hour composite wastewater samples were collected between Sunday and Monday. NIDR data sets were calculated for the following week (from Monday to Sunday) after sampling.

|  | Time from the initiation of IAV wave (weeks) |  |  |  |  |
| --- | --- | --- | --- | --- | --- |
|  | -2 | -1 | 0 | +1 | +2 |
| N of wastewater samples | 4 | 3 | 3 | 4 | 4 |
| N of IAV-positive wastewater samples | 1 | 2 | 1 | 3 | 3 |

## Discussion

The study population consisted of inhabitants in 10 WWTP catchment areas, covering 40% of the Finnish population, and the follow-up was done at 14 months. To the best of our knowledge, this is geographically and temporally the most comprehensive study published to date that tests the effectiveness of IAV monitoring at the national level, which allows for generalisation of the results. During the study period, two waves of an IAV epidemic were monitored in 10 WWTP catchments. Clinical laboratories and doctors are obliged to report the infectious data to the NIDR, allowing both the temporal and spatial correlation of the incidence of IAV in wastewater to the incidence of IAV cases.

The worldwide and local social restrictions because of the SARS-CoV-2 pandemic have changed the temporal prevalence of IAV. The lockdown measures were launched in Finland in March 2020, and they had an immediate impact on the incidence of IAV, which dropped dramatically (Kuitunen, 2021). However, the virus was present in Finland during the study period because a few cases were reported in the NIDR in all months, except in August and September 2021, until the IAV cases increased in November 2021 and a minor peak occurred in December 2021. In the winter months of 2021, incidence was low until April 2021, at which point the incidence increased and the second wave of IAV reached its peak in May 2021. The second wave ended in July, and only a few cases were reported monthly until October 2022.

During the first wave of IAV, infections were reported in 8 out of 10 WWTP catchments, with a median incidence of 2.60. However, the peak incidence was over 5 in only four WWTPs. The IAV was detected in wastewater of seven WWTP catchments, but the virus GC number was so low that most of the samples were under the limit of quantification (LOQ). During the second wave in April and May 2021, more IAV cases were reported, with a mean incidence of 21.30. During the second wave, the virus was detected from all WWTPs, and the virus GC number was over the LOQ.

In the initiation phase of the epidemics, one to two weeks before the first reported cases of IAV, 66.6% of the tested wastewater samples were IAV positive, and one to two weeks after initiation, an additional 71.4% of the samples were IAV positive. These results demonstrate that wastewater surveillance can be useful to detect the virus from the population prior to the first cases being reported in health care facilities, hence providing valuable information about initiation of the IAV epidemics. Earlier, it has been shown that the lead time of wastewater to clinical data was 17 d for the first detection of IAV and three weeks for epidemic detection (Mercier et al., 2022). This is in line with our finding, which had a lower resolution, because of wastewater testing at one-month intervals (min. 3 weeks, max. 6 weeks) and clustering of IAV cases on a weekly basis. The incidence of virus in the population may change rapidly in a period of days (Shaw Stewart and Bach, 2022); therefore, monthly wastewater collection is not optimal for the accurate monitoring of viruses. At the initiation phase of IAV epidemics, the number of infected individuals is low, and even the composite sampling may miss the virus in the influent at the WWTP (Ahmed et al., 2022). In addition, the virus GC number may be close to the detection limit of the assay; therefore, in a time series of wastewater samples, the virus detection may fluctuate between positive and negative (Ahmed et al., 2022). This kind of observation was obtained in two recently published articles, which showed that, in the initiation phase of IAV epidemics, the negative and positive samples fluctuated in daily basis tested wastewater samples (Fig. 2 in Mercier et al., 2022; Fig. 1 in Wolfe et al., 2022). Similar observations can be seen in several surveillance data of the SARS-COV-2 virus in wastewater (Hata et al., 2021; Rusiñol et al., 2021; Tiwari et al., 2022). Therefore, frequent composite wastewater sampling and monitoring of the virus are important for the sensitive and accurate monitoring of IAV epidemics. Ahmed et al. (2022) presented a conceptual model that illustrates the considerations for achieving representative wastewater samples, such as frequent sample collection.

The IAV GC was detected from wastewater in 11 samples without any reported cases of IAV in NIDR during the same week. The lead time between the IAV-positive wastewater sample and reported IAV cases was one to two weeks on seven occasions. An additional four IAV-positive wastewater samples were collected during August and September 2021, when the incidence in Finland was low. The virus GC number was low, and only in one sample was the virus load above LOQ. These results, together with NIDR data, may suggest that IAV was present in Finland all year around. The disease cases in NIDR are underestimated because only the patients with serious symptoms seek medical care and are recognised. Thus, detection of IAV-positive wastewater samples outside the typical IAV season is possible. However, it cannot be excluded that birds may also be one source of these viruses in wastewater. Ducks, geese, gulls and chicken are common in the study locations, and these birds are recognised as the natural reservoirs of avian influenza viruses (Webster et al., 1992). IAVs are shed through faeces from infected birds, and these contaminants contribute to the runoff water to the wastewater network (Markwell and Shortridge, 1982). It can also be speculated about the possibility of the RT-qPCR contamination, but all negative control samples were negative, indicating that contamination was not likely. The RT-qPCR method was tested with the international QCMD panel of influenza A and B virus, and cross-reaction with the influenza B virus was not detected. Reporting of the values below LOQ is important to increase the assay sensitivity and probability of the early detection of epidemics, but the possibility of false positives must be carefully considered.

The estimation of virus load by RT-PCR methods from wastewater can be affected by many environmental and experimental factors, such as sampling, diluted wastewater, RT-qPCR inhibitors and interassay variation of the detection method (Hokajärvi et al., 2021; Zahedi et al., 2021; Alhama et al., 2022; Ahmed et al., 2022). The smoothed average values are one option for interpreting the GC number in wastewater to remove the inaccuracy of the detection assays (Cluzel et al., 2022). In addition, the interfering factors of quantification may be unique for each WWTP catchment. These factors affect the quantification of the virus, and the temporal and spatial comparison of the results may be difficult, even with careful normalisation of the data (Bivins et al., 2021; Cluzel et al., 2022). In the present study, the IAV GC number in wastewater correlated with the incidence of IAV cases in the NIDR. However, the relationship between the IAV GC number in wastewater and incidence rate of disease cases was different between WWTPs. This could be because of the factors mentioned above. The Kendall’s τ correlation strength was over 0.6 (min 0.427, max 0.870) in all except two WWTPs (Table 2), indicating that the strength of the association was similar to an earlier study that showed correlation strength of 0.58 and 0.67 in two locations in the USA (Wolfe et al., 2022). The variation in the correlations between WWTPs may be because of the quite low sample number per WWTP in the current study, but the results show that the surveillance of IAV from wastewater is useful, much like the SARS-CoV-2 surveillance that has been implemented in many countries; hence, its added value for public health may be significant. Based on these and earlier studies (Mercier et al., 2022; Wolfe et al., 2022), WBS can also be applied to the surveillance of IAV when frequent sampling is done.

The present study has shown that the supernatant is a valuable matrix for WBS and that IAV can be detected from it. These results are consistent with a study in which we showed that the SARS-CoV-2 RNA detection and quantification of the copy numbers were only slightly lower in supernatant compared with the solid fraction of freshly collected wastewater samples (Hokajärvi et al., 2021). Two earlier studies reported conflicting data showing that solids are superior to supernatant as a detection matrix for IAV in wastewater. Mercier et al. (2022) presented that, in the primary sludge, IAV was found to be almost exclusively partitioned in solid faction (84.6 ± 10.3% settleable solids and 4.0 ± 3.1% was in suspended solids larger than 0.45 μm), and only a minority of the virus was detected from supernatant (>0,1%). Similar results were shown by Wolfe et al. (2022), who used PEG precipitation for virus concentration. We have shown that the analysis of the solid is important after long-term storge or frozen wastewater samples (Hokajärvi et al., 2021). This is in line with Graham et al. (2021), who showed that SARS-CoV-2 virus was detected mainly from the solid fraction of the wastewater samples frozen prior to the analysis (Graham et al., 2021).

## Limitations of the study

The present study has some limitations. The quality of the clinical data in the Finnish NIDR is good and comprehensive, but the data underestimate the number of clinical cases because only a minority of all IAV cases were laboratory‐confirmed and reported in the NIDR. However, this emphasises that IAV monitoring in wastewater provides useful information and supplements clinical data. Wastewater samples were taken monthly, which limits the accurate temporal comparison with the incidence of reported IAV cases. Despite this limitation, the IAV GC number in wastewater correlated with the incidence cases in NIDR and showed the potential of the WBS for IAV. The RT-qPCR method used for the detection of IAV may also detect zoonotic IAV viruses from wastewater, so it cannot be excluded that zoonotic IAV may have a minor contribution to the results. Still, the strong correlation of the IAV GC number in the wastewater and incidence of IAV cases indicates that the cross-reaction is a minor issue, at least in the current study.

## Conclusion

Poliovirus and, lately, SARS-CoV-2 wastewater surveillance has been implemented in many countries, and its added value for public health is undeniable. The present study confirms that WBS can be applied to influenza A surveillance and could be added to national sentinel surveillance programmes. These results encourage the application of WBS to other epidemic viruses, such as respiratory syncytial virus (RSV) and metapneumovirus. However, it is also important to develop models and approaches that allow holistic use of obtained WBS data for health care needs.

## Supporting information

Supplementary material 1. Preparation of RNA-positive control

## Data Availability

All data produced in the present study are available upon reasonable request to the authors

## Ethical approval

Not required.

## Author statement

The authors have read and approved the revised version.

## Acknowledgements

We want to thank Tampere University laboratory analyst Annika Laaksonen for assistance with the laboratory work and Jussi Lehtonen for the consultation of the statistical methods. We are also grateful to the wastewater treatment plant personnel for collecting and shipping the wastewater samples.

## WastPan Study Group

Viivi Heljanko, Venla Johansson, Paula Kurittu, Ananda Tiwari and Ahmad Al-Mustapha (University of Helsinki); and Anna-Maria Hokajärvi, Anniina Sarekoski, Aleksi Kolehmainen, Teemu Möttönen, Aapo Juutinen, Soile Blomqvist, Kati Räisänen and Carita Savolainen-Kopra (the Finnish Institute for Health and Welfare).

