## Supplementary material 1. Preparation of RNA-positive control for "Wastewater-based surveillance is an efficient monitoring tool for tracking influenza A virus in the community"

Synthetisation of RNA transcripts

The preparation of the RNA-positive control was performed according to Hayes et al. (2020) with minor modifications. Briefly, in the beginning, synthetisation of RNA transcripts of the IAV target sequence was generated commercially by Genscript (Netherlands) on the sequence from GenBank (accession number MW855999). The 106 bp long DNA string of IAV included the priming sites for the primers and probes of the IAV (Gene Matrix) target sequences (WHO, 2021). The DNA string was cloned into a pET3a plasmid vector using Genscript (Netherlands). The plasmid vector was transformed into DH5 alpha *E. coli* cells by electroporation (Gene pulser^®^ II RF Module, BioRad, Cat. 165-2105), the transformed *E. coli* was grown on ampicillin-resistant Luria Bertani agar plates (LB) at 37 ^o^C for overnight, and single colonies were picked and grown in LB broth medium for plasmid extraction using the GeneJET plasmid Midi-prep kit (Thermo Fisher Scientific Baltics UAB, Lithuania), per the manufacturer’s instructions. The sequence of the target molecule from several colonies was verified by PCR analysis with M13 primers (Table S1) of the vector, and the amplicons were run by 1% agarose gel electrophoresis. Furthermore, the PCR products were sequenced to confirm the sequence of the clones (Macrogen, Netherlands). The concentration and volume of the reagents and the PCR programme used are described in Tables S2 and S3. Plasmid was linearised at the 3’end with ClaI (Promega, Madison, WI, USA, 10 u/µL) digestion as per the manufacturer’s instructions. RNA transcript was produced using MEGAscripts T7 transcription kit (ThermoFisher Scientific Baltics UAB, Lithuania), followed by removal of DNA with DNase treatment.

Purification and quantification of RNA

Transcribed RNA was purified using an RNeasy mini kit (Qiagen, Germany) and quantified using a NanoDrop One Microvolume UV VIS spectrophotometer (Thermo Fisher Scientific). The concentrations obtained from Nanodrop were used to calculate the copy numbers of the transcript products, presuming 330g/mol molecular mass of each base. A tenfold serial dilution (10^5^-10^1^ copies/ µL) was made from RNA transcripts using RNA storage solution (ThermoFisher Scientific Baltics UAB, Lithuania, pH 6.4) and RNase inhibitor (ThermoFisher Scientific Baltics UAB, Lithuania, 20 U/µL) to obtain a long preservative life. The dilution series of RNA transcripts was preserved in small aliquots at -80 ^°^C. The standard curves of tenfold serial dilution of control materials were tested using IAV-specific RT-PCR described in the article.

Table S1. Sequences of M13 universal primers.

| Primers | Sequence (5’-3’) |
| --- | --- |
| M13 forward | TGTAAAACGACGGCCAGT |
| M13 Reverse | GGAAACAGCTATGACCATG |

Table S2. Concentrations and volumes of reagents in the detection of transformed products by M13 primers.

| Component | 25 μl reaction | Final concentration |
| --- | --- | --- |
| 10X Standard *Taq* Reaction Buffer | 2.5 μl | 1X |
| 10 mM dNTPs | 0.5 µl | 200 µM |
| 10 µM M13 forward Primer | 0.5 µl | 0.2 µM |
| 10 µM M13 Reverse Primer | 0.5 µl | 0.2 µM |
| Template DNA | 0.5 µl | variable |
| *Taq* DNA Polymerase | 0.125 µl | 0.625 units |
| Nuclease-free water | 21.0 µl |  |

Table S3. PCR programme used for M13 run during preparation of positive control.

| Temperature | Time | Cycles |
| --- | --- | --- |
| 95 °C | 3 min. | 1x |
| 95 °C | 45 sec. | 35x |
| 55 °C | 45 sec. |  |
| 72 °C | 5 min. |  |
| 72 °C | 5 min. | 1x |
| 4 °C | Infinite |  |

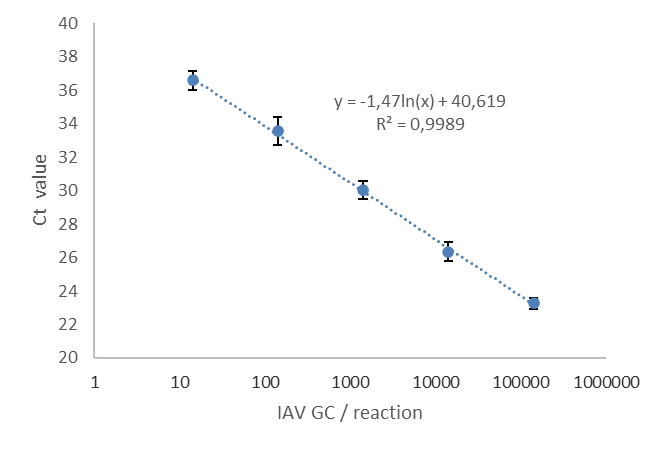

Figure S1. Quantitative control series of the IAV RT-qPCR method. The standard curve was calculated for all control series. The average amplification efficacy was 1.974 (min. 1.861 and max. 2.094), and the average of the standard curve slope was 3.381 (min. 3.115 and max. 3.708).
